# Frontline maintenance olaparib therapy for BRCA/HRD-positive advanced epithelial ovarian cancer: real-world data analysis in matched population

**DOI:** 10.1101/2024.10.07.24315067

**Authors:** Alexey A. Rumyantsev, Elena V. Glazkova, Tatiana E. Tikhomirova, Ilya A. Pokataev, Ekaterina O. Ignatova, Rostislav I. Knyazev, Alexandra S. Tyulyandina, Sergey A. Tjulandin

## Abstract

**Background:** introduction of the poly(ADP-ribose) polymerase (PARP) inhibitors to clinical practice remarkably improved outcomes for advanced epithelial ovarian cancer (EOC) patients. We conducted this study to evaluate efficacy of frontline maintenance olaparib therapy in BRCA-mutated and/or HRD-positive EOC patients in real-world practice setting.

**Patients and methods:** we enrolled patients with FIGO stage III-IV high-grade serous or endometrioid EOC with BRCA1/2 mutations and/or HRD-positive status with complete or partial response to frontline therapy, who were treated in 2014-2024. Main objective of this trial was to compare progression-free survival (PFS) of HRD+/BRCA-mutant advanced EOC patients treated with or without maintenance olaparib in well-balanced treatment arms. Cardinality matching was considered to ensure balancing of the study arms with 1:1 ratio of patients in trial arms. The groups were balanced according to the presence of residual tumor after initial treatment, platinum-free interval duration after frontline therapy, secondary local therapy for recurrent disease, treatment with platinum drugs for relapse and subsequent bevacizumab. The primary endpoint of the study was PFS.

**Results:** cardinality matching with 1:1 ratio resulted in 282 matched patients for the analysis. Groups were well balanced in all baseline characteristics. Median age in both treatment arms was 50 years with no differences in patients’ age, surgical outcomes, prevalence of BRCA-mutated or HRD-positive disease and response to initial platinum-based therapy. With a median follow up of 37.2 mo. median PFS was 38.0 in the olaparib arm and 13.7 mo. in the control arm, respectively (HR 0.32; 95% CI 0.23-0.43; p<0.001). Estimated 3-year PFS was 50.9% and 11.5%, respectively. Median PFS2 was 49.5 mo. in the olaparib arm compared to 34.3 mo. in the control arm (HR 0.58; 95% CI 0.39-0.85; p=0.005).

**Conclusion:** this study confirms the benefits of olaparib maintenance therapy in patients with HRD- -positive and/or BRCA-mutated advanced EOC in real-world setting.

**Highlights:**

- Using cardinality matching two cohorts of patients after frontline therapy were made (treated with maintenance olaparib vs not);
- Median PFS was 38.0 in the olaparib arm and 13.7 mo. in the control arm, respectively (HR 0.32; 95% CI 0.23-0.43; p<0.001);
- This data confirms efficacy of maintenance olaparib for BRCA/HRD-positive advanced epithelial ovarian cancer in routine clinical practice.

## Introduction

Epithelial ovarian cancer (EOC) is a potentially life-threatening disease which tends to recur after primary treatment. Development and introduction of the poly(ADP-ribose) polymerase (PARP) inhibitors to clinical practice remarkably improved outcomes for patients with advanced EOC, especially for those with tumors harboring BRCA1/2 mutations or other genomic alterations leading to homologous recombination deficiency (HRD). SOLO1 (n=391) trial has shown 67% reduction in risk of diseases progression or death of BRCA-mutated EOC with corresponding median progression-free survival (PFS) of 56.0 and 13.8 mo. in patients treated with olaparib and placebo respectively ^1^. There was a clinically meaningful improvement in overall survival (OS), at 7 years 67.0% and 46.5% of patients in olaparib and placebo arm were alive (hazard ratio [HR] 0.55; 95% CI 0.40-0.76; p=0,0004), albeit not statistically significant according to statistical hypothesis threshold for the analysis (p<0,0001) ^2^.

Several other trials confirmed efficacy of PARP inhibitors is initial EOC treatment. PAOLA-1 trial (n=806) has shown significant improvement of PFS for HRD-positive EOC patients treated with olaparib and bevacizumab combination compared to placebo and bevacizumab (HR 0.41; 95% CI 0.32-0.54). This translated in OS benefit for HRD-positive patients, at 5 years 65.5% and 48.4% in olaparib and placebo arms were still alive (HR 0.62; 95% CI 0.45-0.85) ^3,4^. PRIMA trial (n=733) demonstrated efficacy in even broader patient population with PFS improvement from 8.2 mo. to 13.8 mo. with maintenance niraparib therapy compared to placebo (HR 0,43; 95% CI 0.50-0.76; p<0.001) in unselected advanced EOC population. Expectedly, compared to HRD-positive (HR 0.52; 95% CI 0.40-0.68) populations, patients with HR-proficient (HRp) tumors had little clinical benefit from therapy with 3 mo. absolute improvement in median PFS (HR 0.65; 95% CI 0.49-0.87) ^5,6^. Collectively, this data made maintenance PARP inhibitors a standard of care for BRCA-mutated or HRD-positive EOC ^7,8^.

However, eligibility criteria may restrict diverse patient enrollment in clinical trials of therapeutic agents and clinical trial results do not always correspond to real-world practice and patient outcomes. There is growing interest in using real-world data to assess reproducibility of trials data in clinical practice setting^9^. We conducted this retrospective multicenter study to evaluate efficacy of frontline maintenance olaparib therapy in BRCA-mutated and/or HRD-positive EOC patients in real-world practice setting.

### Patients and methods

For this retrospective study we enrolled patients with FIGO stage III-IV high-grade serous or endometrioid advanced epithelial ovarian cancer (EOC) with pathogenic BRCA1/2 mutations and/or HRD-positive status (defined by AmoyDx HRD Focus Panel ^10^), who were treated in 2014-2024 years. Patients were selected from N.N. Blokhin NMRCO ovarian cancer database. Additional inclusion criteria were complete response (or no evidence of disease (NED) for patients without residual disease after surgery) to frontline therapy. Patients with stable disease or progressive disease during frontline therapy were excluded from the analysis. Mutation status (BRCA1/2 and/or HRD-status) were assessed before start of maintenance therapy. Consistently with the trial design, we included patients who were treated with at least 1 dose of maintenance olaparib after completion of the primary treatment in olaparib arm included. Per the decision of the treating physician patients might be treated with bevacizumab or endocrine therapy, however, use of investigational anticancer agents was not allowed. Patients in the control arm could be treated with PARP-inhibitors in subsequent lines of anticancer treatment.

The primary objective of this retrospective trial was to compare progression-free survival (PFS) of HRD+/BRCA-mutant advanced EOC patients treated with or without maintenance olaparib in well-balanced treatment arms. Cardinality matching was considered to ensure adequate balancing of the study arms. Cardinality matching is a method for finding the largest possible number of matched pairs of exposed and unexposed individuals from an observational dataset ^11^. The covariates considered for the matching were age, residual disease after surgery (absent or present), timing of surgery (primary debulking surgery vs interval), clinical risk group according to PAOLA-1 trial criteria (high vs low risk), tumor genetic status (presence of BRCA1 vs BRCA2 mutations vs BRCAwt/HRD+), FIGO stage of the disease (III vs IV) and response to primary treatment (partial response (PR) vs complete response (CR) or no evidence of disease (NED) after completion of treatment).

Response to treatment was assessed by treating physicians before olaparib administration and by the investigators using medical charts before inclusion in this trial. Clinical CR or NED status was defined as no evidence of measurable or non-measurable disease on the post-treatment scans per RECIST1.1 criteria and a normal CA-125 levels either after complete debulking surgery or CR to anticancer treatment. PR was defined as ≥30% reduction in size of measurable lesions demonstrated from the start to finish of chemotherapy OR no evidence of RECIST measurable disease on the post-treatment scan with a CA-125 which has not decreased to within the normal range

For propensity scores estimation for cardinality matching, the mentioned covariates were used as main effects in a logistic regression analysis, where treatment with olaparib was a dependent variable (outcome) and the covariates were treated as independent variables. Several separate models were fitted and modeled the probability of a patient in the analysis population being treated with or without maintenance olaparib. The decision on the final set of covariates included in the primary matching model was based on several factors, including model fit statistics, balancing of the covariates, amount of missing data and final sample size after matching. Decisions were made without knowledge of the impact on the outcome of the survival analyses. All initially planned covariates were included in the cardinality matching model. Age was dichotomized to two categories (<60 years vs ≥60 years) as well as tumor genetics (BRCA1/2), as this allowed to achieve the best fir of the model.

Patients received Maintenance therapy with olaparib per the approved label (300 mg BID for tablet and 400 mg BID for capsule formulation). Dose reduction, treatment delays and interruptions, and prescription of additional supportive therapy were made per the decision of treating physician and label. Disease assessments were done per local practice and included computed or magnetic resonance tomography with contrast enhancement and CA-125 measurement every 9-12 weeks for 2 years after treatment completion and every 12-18 weeks thereafter.

The primary endpoint of this trial was PFS which was calculated from the date of primary therapy completion to disease progression or death due to any cause. PFS2 was calculated from the date of primary therapy completion to subsequent disease progression or death due to any cause. Median follow up time was assessed using reverse Kaplan-Meier method. Sensitivity analysis were preplanned to assess PFS adjusted for potential residual imbalances in key prognostic factors, initially used in cardinality matching, impact of exclusion of bevacizumab and BRCA wild-type (wt)/HRD-positive patients from analysis.

All analyses were performed using R Statistical Software (v4.1.2; R Core Team 2021) and RStudio (v. 200.07.2.576, RStudio Team 2022). Cardinality matching was conducted with MatchIt package^11^.

## Results

Overall, 394 patients with advanced epithelial ovarian cancer were selected for this retrospective study (Figure 1 depicts flowchart for patients selection from the database), of whom 179 received maintenance olaparib after frontline chemotherapy and 189 did not. Table 1 summarizes patient characteristics in the initial data set and after cardinality matching analysis. In the initial data set (n=394) there were significant imbalances in characteristics in terms of proportion of patients with HRD+/BRCAwt tumors in olaparib vs control arm (15.6% vs 4.65%, respectively; p<0.001), without residual tumor after either primary or interval debulking surgery (43.1% vs 21.4%) as well as complete response or no evidence of disease after completion of frontline therapy (87.7% vs 62.4%; p<.001). Further, there was trend for older age in the olaparib arm, as 26,8% in this arm was >60 years old compared to 19.0% in the control arm (p=.067).

**Table 1.** Patient characteristics.

|  | All patients |  |  | Matched population |  |  |
| --- | --- | --- | --- | --- | --- | --- |
|  | Olaparib | Control | p | Olaparib | Control | p |
| N | 179 | 185 |  | 141 | 141 |  |
| Age, median | 51 (34-82) | 50 (32-77) | .245 | 50 (34-82) | 50 (32-77) | .676 |
| ≤60 years | 131<br>(73.2%) | 174<br>(80.9%) | .067 | 110 (78.0%) | 110 (78.0%) | 1.000 |
| >60 years | 48 (26.8%) | 41<br>(19.0%) |  | 31 (22.0%) | 31 (22.0%) |  |
| FIGO IV | 39 (21.8%) | 38<br>(17.7%) | .306 | 23 (16.3%) | 21 (14.9%) | .744 |
| HGSOC | 179<br>(100%) | 185<br>(100%) | 1.000 | 141 (100%) | 141 (100%) | 1.000 |
| **BRCA1/2mut** | **151**<br>**(84.4%)** | **205**<br>**(95.3%)** | **<.001** | **129 (91.5%)** | **131 (92.9%)** | **.658** |
| **HRD+/BRCAwt** | **28**<br>**(15.6%)** | **10**<br>**(4.65%)** |  | **12 (8.51%)** | **10 (7.1%)** |  |
| PDS | 107<br>(60.0%) | 129<br>(60.0%) | .964 | 92 (65.2%) | 95 (67.3%) | .707 |
| **No residual tumor** | **77**<br>**(43.1%)** | **46**<br>**(21.4%)** | **<.001** | **44 (31.2%)** | **41 (29.1%)** | **.698** |
| High risk* | 137<br>(76.5%) | 180<br>(83.7%) | .074 | 109 (77.3%) | 111 (78.7%) | .775 |
| Bevacizumab | 15 (8.38%) | 14<br>(6.51%) | .481 | 11 (7.80%) | 11 (7.80%) | 1.000 |
| **CR/NED** | **157**<br>**(87.7%)** | **134**<br>**(62.4%)** | **<.001** | **119 (84.4%)** | **117 (83.0%)** | **.748** |
| Subsequent PARPi | - | 133<br>(61.9%) | - | - | 94 (66.7%) | - |

HGSOC – high-grade ovarian carcinoma; PDS – primary debulking surgery; CR/NED – complete response or no evidence of disease; \*per PAOLA-1 trial criteria; PARPi – PARP-inhibitors

**Figure 1.**
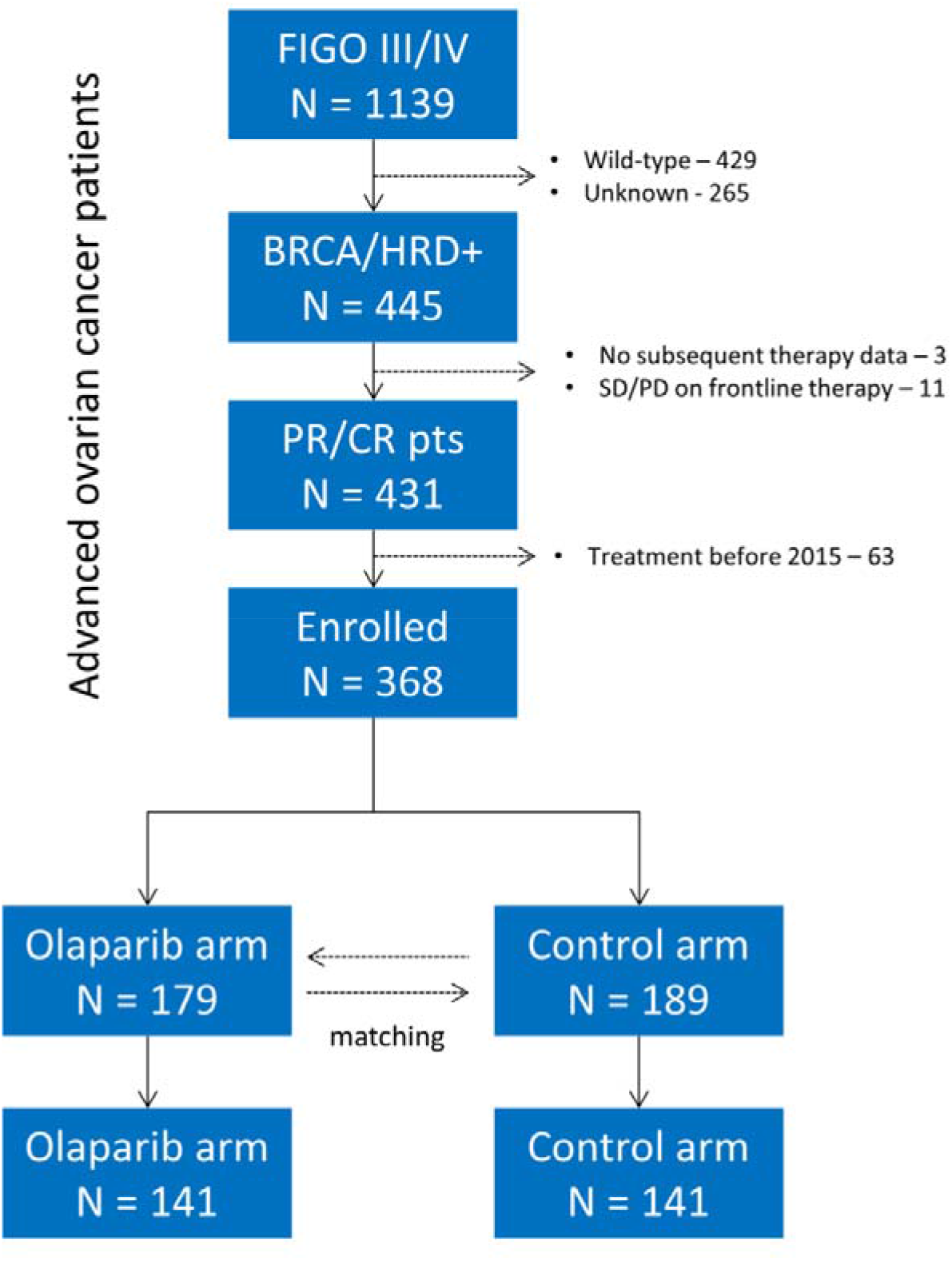
Patients flowchart in the initial dataset and matched population.

Cardinality matching with 1:1 ratio resulted in 282 matched patients for the further analysis. Both groups were well balanced in all baseline characteristics, including the previously mentioned variables, with statistical significant imbalances. Median age in both treatment arms was 50 years with no differences in amount of patient age >60, accounting for 31 (22%) in both arms (p=1.000), 12 (8.51%) and 10 (7.1%) patients had HRD+/BRCAwt tumors, 97 (68.8%) and 100 (70.9%) of patients had no residual tumor after debulking surgery (p=0.698) and 119 (84.4%) and 117 (83.0%) had complete response or NED after completion of frontline treatment (p=0.748). Standardized mean differences are shown on Figure 2. Of note, 7 patients with HRD+/BRCAwt tumors received off-label olaparib therapy without concomitant bevacizumab, 4 patients received olaparib and bevacizumab combination.

**Figure 2.**
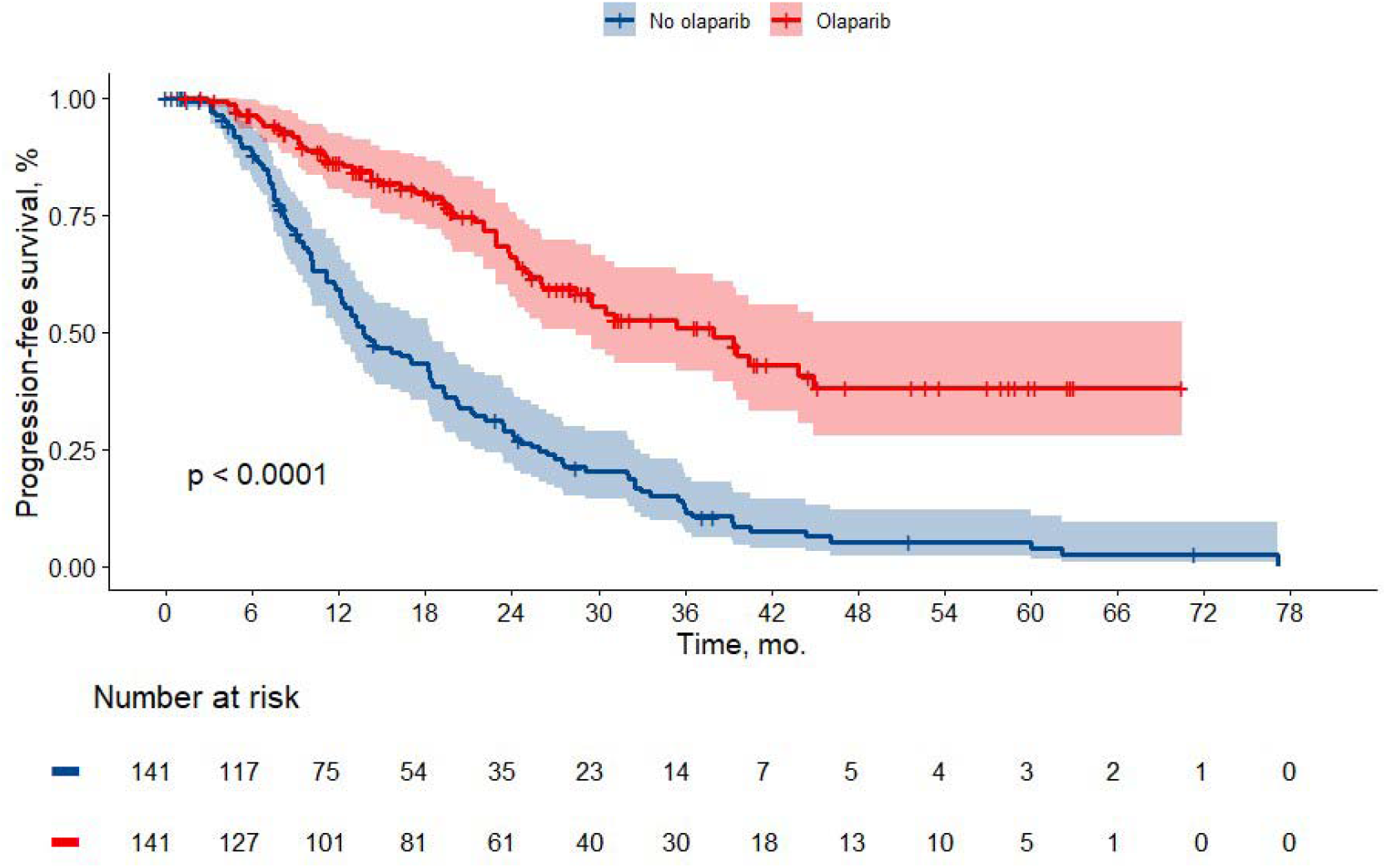
Kaplan-Meier estimates of progression-free survival (PFS) in olaparib and control arms in the matched population

With a median follow up of 37.2 mo. (31.2 mo. in the olaparib arm and 61.0 mo. in the control arm, respectively) median PFS was 38,0 in the olaparib arm and 13.7 mo. in the control arm, respectively (HR 0.32; 95% CI 0.230-0.43; p<0.001). Estimated 3-year PFS was 50.9% and 11.5%, respectively (Figure 2). The differences were clinically meaningful and statistically significant. As there was imbalance in median follow up time between trial arms restricted means survival time (RMST) analysis was conducted to further evaluate efficacy of olaparib. Consistently with the primary analysis, 3-year RMST for PFS in olaparib arm and control arms was 27.6 mo. and 17.5 mo., respectively with significant corresponding between-group contrast 10.1 mo. (95% CI 7.39-12.7). Median PFS2 was 49.5 mo. in the olaparib arm compared to 34.3 mo. in the control arm (HR 0.58; 95% CI 0.39-0.85; p=0.005), Kaplan-Meier estimates are in the Figure 3.

**Figure 3.**
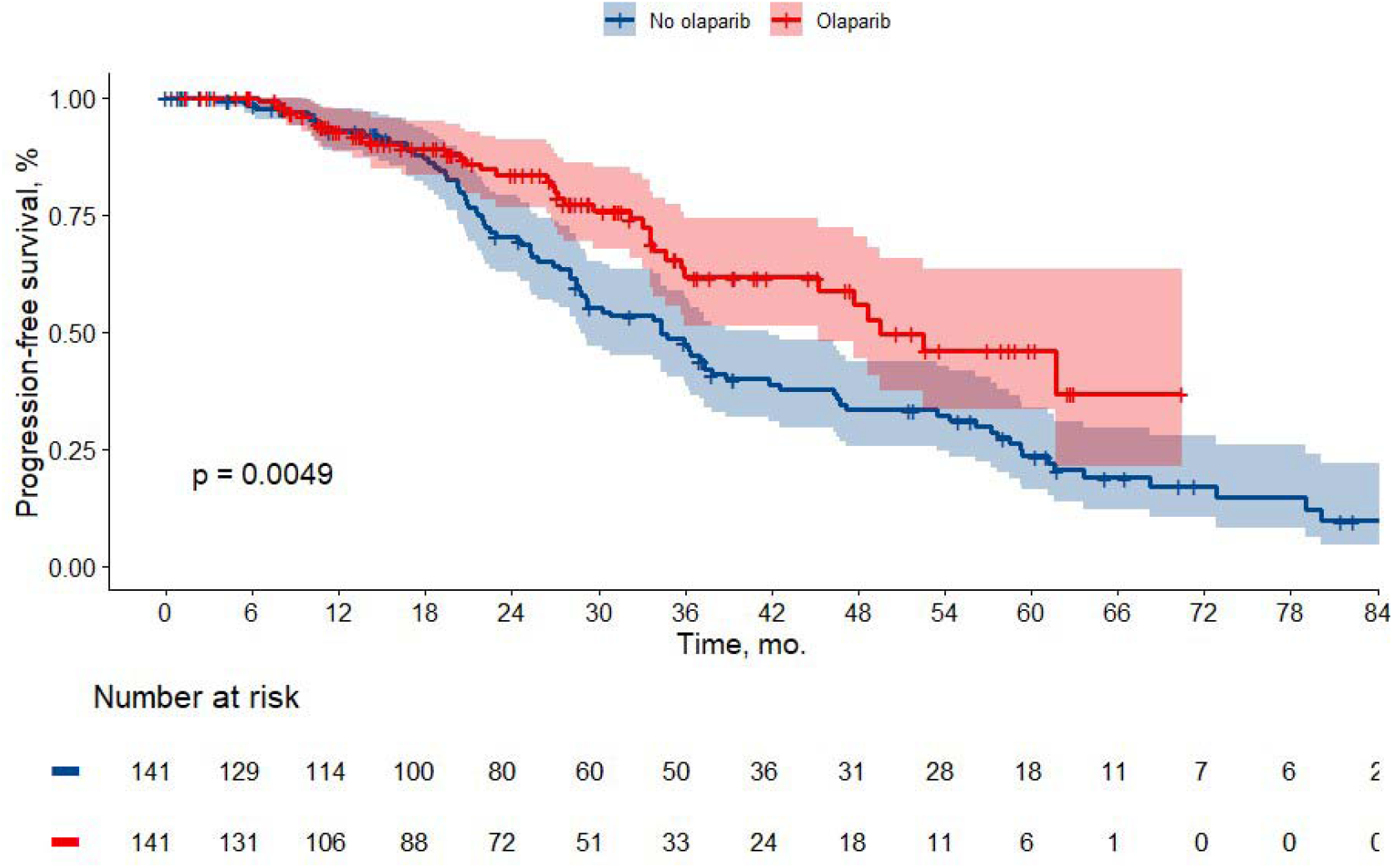
Kaplan-Meier estimates of progression-free survival 2 (PFS2) in olaparib and control arms in the matched population

Additional sensitivity analyses were conducted. Neither adjustment for key prognostic factors in multivariate Cox regression (HR 0.32; 95% CI 0.23-0.44), nor exclusion of patients with BRCAwt/HRD+ tumors or those treated with bevacizumab had significant impact on olaparib efficacy. The subgroup analysis of PFS according to baseline characteristics showed no evidence for heterogeneity of treatment effect size (Figure 4).

**Figure 4.**
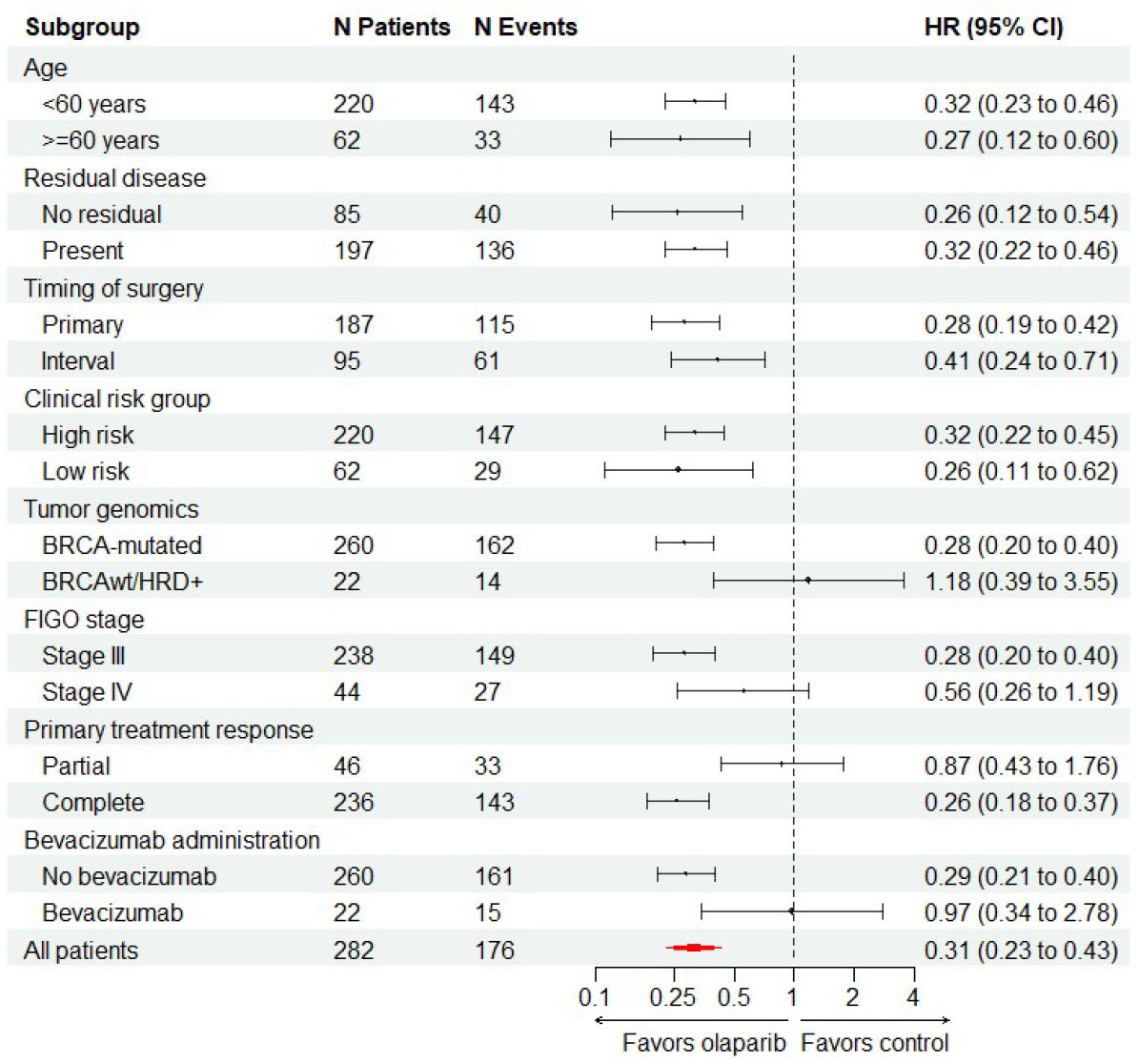
Forest plot of the treatment effect on PFS in subgroup analyses.

## Discussion

The results of our trial indicate significant improvement in outcomes of BRCA-mutated and/or HRD-positive in matched EOC patient population. Hazard ratio of disease progression with olaparib maintenance therapy was reduced by 68%, with corresponding 3-year PFS estimated 3-year PFS was 50.9% in the olaparib and 11.5% in the control arm, respectively. Our findings are consistent with the primary analysis results of SOLO1 trial where with median follow-up of 41 mo. 70% reduction in hazard ratio of disease progression or death. At the same time, 3-year PFS in SOLO1 was 60% and 27% in olaparib and placebo arm respectively, however this seems to be related to prevalence of the low clinical risk group in the latter trial, as 77% of patients had no residual disease in SOLO1 compared to only 35% in our trial population.

Eligibility criteria for this trial largely reflect criteria for SOLO1 trial, as we enrolled patients with the same histologic subtypes, stages of EOC and most of the patients in our trial had BRCA1/2-mutated disease. We measured PFS from the end of frontline therapy in an attempt to achieve better precision in survival comparison with SOLO1. We also enrolled patients treated with bevacizumab and HRD-positive tumors not harboring BRCA mutation, however the number of these patients in the final trial dataset appears to be relatively small and unlikely to affect the results. Hence, the results of our real-world data study confirms the initial results of SOLO1 and claim the same benefits for advanced EOC patients.

More patients in the olaparib arm in our trial were censored in the survival analysis due to shorter follow-up time. This finding may be easily explained by introduction of PARP inhibitors in routine clinical practice of frontline EOC treatment in 2019-2020, however this may lead to instable survival estimates and significantly inflate type I error in the analysis. However, early separation in the survival curves was observed with significant PFS improvement of patients treated with olaparib after 12 month of treatment. Furthermore, sensitivity analyses with random patient censoring in the control arm and event occurrence in the olaparib arm did not impact clinical or statistical significance of frontline olaparib therapy in advanced ovarian cancer patients.

Interestingly, in HRD-positive patients without pathogenic mutations in BRCA1/2 we found no differences in survival of patients (HR 1.18; 95% CI 0.39-3.55), however, this should be interpreted with extreme caution due to low number of patients in this subgroup. Wide confidence interval does not allow ruling out neither benefit nor detrimental effects in this patient population.

We did not attempted to assess overall survival of patients treated with olaparib due to relatively short follow up time in our trial. As was previously noted in SOLO-1 trial, separation of the survival curves occurred only 5 years after randomization and clinically meaningful differences were achieved after 7 years of follow-up^2^. However, our data indicates similar benefits in terms of PFS and PFS2, which might improve OS over the following years. Significant differences in PFS were achieved despite high cross-over rate, as many as 66.7% of patients from the control arm received subsequent therapy with PARP-inhibitors.

This was a retrospective study, which are subject to various inherited biases, potential influence of a number of unidentified factors that have significant impact on the results cannot be excluded.. Cardinality matching which was implemented for arm balancing in this trial may somehow alleviate such imbalances and our study was conducted in well matched cohort of patients. Nonetheless, the magnitude of benefit of maintenance olaparib therapy in this study is largely equal to reported in SOLO1 and PAOLA-1 trials which support routine administration of this treatment.

## Conclusion

Our trial has shown improved outcomes in HRD/BRCA-positive advanced epithelial ovarian cancer with maintenance olaparib after frontline treatment in real clinical practice setting.

## Data Availability

Data produced in the present study are available upon reasonable request to the authors

## Conflict of Interest Statement

The authors declare following conflicts of interest:

- Rumyantsev A: Financial Interests, Personal, Speaker’s Bureau: Biocad, AstraZeneca, Pfizer, MSD, Merck.
- E. Glazkova: Financial Interests, Personal, Speaker’s Bureau: Biocad, AstraZeneca, R-Pharm, MSD, Pfizer.
- T. Tikhomirova: Speaker’s Bureau: AstraZeneca
- Pokataev: Financial Interests, Personal, Speaker’s Bureau: AstraZeneca, Johnson & Johnson, MSD, Biocad, Sanofi, Roche, Eli Lilly.
- E.Ignatova: declares no conflict of interest
- R. Knyazev: declares no conflict of interest
- A. Tyulyandina: Financial Interests, Personal, Speaker’s Bureau: AstraZeneca, BIOCAD, Merck, Pfizer, Takeda, MSD. Employment: AstraZeneca.
- S. Tjulandin: Financial Interests, Personal, Ownership Interest: RosPharmTech; Financial Interests, Personal, Speaker’s Bureau: AstraZeneca, Biocad, Eli Lilly, MSD.

## Funding Information

This study received no funding.

## Notes

### Competing Interest Statement

Rumyantsev A: Biocad; AstraZeneca; Pfizer; MSD; Merck.
E. Glazkova: Biocad; AstraZeneca; R-Pharm; MSD; Pfizer.
T. Tikhomirova: AstraZeneca
Pokataev: AstraZeneca; Johnson & Johnson; MSD; Biocad; Sanofi; Roche; Eli Lilly.
E.Ignatova: declares no conflict of interest
R. Knyazev: declares no conflict of interest
A. Tyulyandina: AstraZeneca; BIOCAD; Merck; Pfizer; Takeda; MSD. Employment: AstraZeneca.
S. Tjulandin: RosPharmTech; AstraZeneca; Biocad; Eli Lilly; MSD.

### Author Declarations

N.N. Blokhin NMCO IRB waived ethical approval for this work as only deidentified patient data was used

## References

1. Banerjee S, Moore KN, Colombo N, et al. Maintenance olaparib for patients with newly diagnosed advanced ovarian cancer and a BRCA mutation (SOLO1/GOG 3004): 5-year follow-up of a randomised, double-blind, placebo-controlled, phase 3 trial. Lancet Oncol. 2021;22(12):1721–1731. doi:10.1016/S1470-2045(21)00531-3

2. DiSilvestro P, Banerjee S, Colombo N, et al. Overall Survival With Maintenance Olaparib at a 7-Year Follow-Up in Patients With Newly Diagnosed Advanced Ovarian Cancer and a BRCA Mutation: The SOLO1/GOG 3004 Trial. J Clin Oncol Off J Am Soc Clin Oncol. 2023;41(3):609–617. doi:10.1200/JCO.22.01549

3. Ray-Coquard I, Leary A, Pignata S, et al. Olaparib plus bevacizumab first-line maintenance in ovarian cancer: final overall survival results from the PAOLA-1/ENGOT-ov25 trial. Ann Oncol. 2023;34(8):681–692. doi:10.1016/j.annonc.2023.05.005

4. Ray-Coquard I, Pautier P, Pignata S, et al. Olaparib plus Bevacizumab as First-Line Maintenance in Ovarian Cancer. N Engl J Med. 2019;381(25):2416–2428. doi:10.1056/NEJMoa1911361

5. González-Martín A, Pothuri B, Vergote I, et al. Progression-free survival and safety at 3.5 years of follow-up: results from the randomised phase 3 PRIMA/ENGOT-OV26/GOG-3012 trial of niraparib maintenance treatment in patients with newly diagnosed ovarian cancer. Eur J Cancer. 2023;189:112908. doi:10.1016/j.ejca.2023.04.024

6. González-Martín A, Pothuri B, Vergote I, et al. Niraparib in Patients with Newly Diagnosed Advanced Ovarian Cancer. N Engl J Med. 2019;381(25):2391–2402. doi:10.1056/NEJMoa1910962

7. González-Martín A, Harter P, Leary A, et al. Newly diagnosed and relapsed epithelial ovarian cancer: ESMO Clinical Practice Guideline for diagnosis, treatment and follow-up. Ann Oncol. 2023;34(10):833–848. doi:10.1016/j.annonc.2023.07.011

8. NCCN Clinical Practice Guidelines in Oncology (NCCN Guidelines®). Ovarian Cancer, including Fallopian Tube Cancer and Primary Peritoneal Carcinoma. Version 3.2024. Published July 15, 2024. Accessed August 5, 2024. https://www.nccn.org/professionals/physician_gls/pdf/ovarian.pdf

9. Sherman RE, Anderson SA, Dal Pan GJ, et al. Real-World Evidence — What Is It and What Can It Tell Us? N Engl J Med. 2016;375(23):2293–2297. doi:10.1056/NEJMsb1609216

10. Kekeeva T, Andreeva Y, Tanas A, et al. HRD Testing of Ovarian Cancer in Routine Practice: What Are We Dealing With? Int J Mol Sci. 2023;24(13):10497. doi:10.3390/ijms241310497

11. Ho DE, Imai K, King G, Stuart EA. MatchItD: Nonparametric Preprocessing for Parametric Causal Inference. J Stat Softw. 2011;42(8). doi:10.18637/jss.v042.i08

